# ASSOCIATION BETWEEN THE USE OF CANNABIS AND DEPRESSION IN UNITED STATES ADULTS

**DOI:** 10.1101/2023.10.25.23297562

**Authors:** K. Ortiz, J. OLeary, H. Bhatt, V. Nadraga

## Abstract

**Introduction:** Recent studies show a growing body of evidence suggesting that users of cannabis have consistently higher prevalence rates of depressive disorders in comparison to non-users. Besides, it can be used as a highly effective treatment for depression and other mood disorders.

**Methods:** A study with a cross-sectional design was conducted with data from respondents aged 25-64 who participated in the 2018 Behavioral Risk Factor Surveillance System (BRFSS) survey. The population was assessed for the baseline characteristics, followed by a bivariate analysis, a multivariate logistic regression to control any confounders, and assess the association between the use of cannabis and depression.

**Results:** The sample included 57,757 individuals. The unadjusted binary logistic regression indicated that those who use cannabis are 93% more likely (OR: 1.93) to have a diagnosis of depression. On the other hand, the adjusted analysis indicated that those who use cannabis are 78% more likely (OR: 1.78) to have a diagnosis of depression. Other variables including participants under the age of 45 years were significantly associated with the diagnosis of depression (OR: 1.35). Moreover, participants with an education without a high school diploma (OR: 0.54), and those having a full-time commitment (OR: 0.37) were significantly less likely to have a diagnosis of depression.

**Conclusion:** Findings suggest that selected U.S. participants who use cannabis have an increased risk to report depression. Furthermore, factors such as age, level of education, and a person’s time commitment status were found to have a significant influence on whether a diagnosis of depression is present.

## Introduction and Background

Depressive disorder was the third overall source of the burden of disease around the world in 2008 (1), according to the World Health Organization (WHO). At the beginning of 2023, the Institute of Health Metrics and Evaluation estimated approximately 280 million people in the world have depression (2). However, by the year 2030, the WHO has projected that Major Depression Disorders (MDD) will be ranked first (1). Known as one of the most common, highly prevalent, and disabling of the mood disorders, the disease tends to cause a persistent feeling of sadness and anhedonia, the loss of ability to feel pleasure; it can also be diagnosed when an individual has constant feelings of worthlessness or guilt, poor concentration, psychomotor agitation, lack of energy and sleep disturbances, or even suicidal thoughts (1). To be diagnosed with this debilitating disease, as per the Diagnostic and Statistical Manual of Mental Disorders 5 Edition (DSM-5), an individual must be expressing at least five of the above-mentioned symptoms, of which one must be anhedonia or depressed mood causing impairment socially or occupationally (3). The etiology has been proposed to be of multifactorial origin, including genetic, biological, psychosocial, as well as environmental factors (4). It was earlier believed to be primarily due to irregularities of naturally produced neurotransmitters, especially dopamine, serotonin, and norepinephrine; and this theory has been backed up over the years using antidepressants that inhibit the reuptake of these neurotransmitters, leading to higher levels being retained and helping in the treatment of depression (4).

According of the NIH (National Center for Complementary and Integrative Health), the cannabis, commonly known as marijuana, is a cannabinoid drug found in the cannabis sativa plant, it contains psychoactive cannabinoid THC (delta-9-tetrahydrocannabinol) and the non-psychoactive cannabidiol (CBD) (5). In recent studies, there is a growing body of evidence suggesting that users of cannabis have consistently higher prevalence rates of depressive disorders in comparison to nonusers (6). However, those have also demonstrated that cannabis can be used as a highly effective treatment for depression and other mood disorders (3). With this contradiction in mind, the aim of this study is to assess the association between the use of cannabis and depression.

Cannabis use is one of the most consumed illicit substances among different age groups, genders, and race. The prevalence of cannabis use is staggering, with upwards of 205 million individuals having consumed it at least once in 2009 alone (7). At the same time, depending on age range and cultural background, it is estimated that some demographics have upwards of 50% of individuals having tried the drug at least 1 time (7). Concurrently, illicit use of cannabis has many adverse effects on the human body, affecting almost every major body system (3). However, it is this illicit use of the substance that has cast a negative shadow over its use both illegally and for medical purposes. In an extensive research project over a 40-year span, Onaemo et al. have shown that the comorbidity of cannabis use results in major depression and general anxiety disorder have negative externalities (8). The overall application of this research will provide educational material exhibiting the cannabis use in relation of having a diagnosed of depression. In addition, medical providers could be able to use this research to educate themselves about the effects of cannabis use as the drug is gaining widespread legalization and general acceptance.

## Methodology

We performed a cross-sectional study based on secondary analysis of data from the Behavioral Risk Factor Surveillance System (BRFSS) questionnaire from 2018 (9). The BRFSS is a joint project, which started in 1984, between all the participating states and territories in the US and the Centers for Disease Control and Prevention (CDC) (10). It has been an apparatus of uninterrupted health-related surveys over the phone which helps accumulate data on state-specific chronic diseases and comorbidities, access to health care, use of preventive health services, and health risk behaviors, all related to the leading causes of death and disability in noninstitutionalized adult (over the age of 18) residents of the U.S.; as of 2018, the data collected was inclusive of all 50 states, the District of Columbia, Guam, and Puerto Rico (10).

The population of our study consisted of participants of the 2018 BRFSS survey. As criteria for inclusion was age between 25 and 64 years old and if they answered the following question: (Ever told) you have a depressive disorder (including depression, major depression, dysthymia, or minor depression)? with either “yes” or “no”. As criteria for exclusion was if they report more than 14 alcoholic drinks per week or 60 alcoholic drinks per month, or any use of tobacco. The independent variable, the status of cannabis use defined according to the question “During the past 30 days, on how many days did you use marijuana or cannabis?” Based on the response options (01-30 Number of days, None, don’t know/not sure, Refused), the participants will be categorized in two groups who either use cannabis either 1 day or more (group 1), or don’t use it at all (group 2). If the response is “Don’t know/not sure or Refused” they will be excluded from the study. The dependent variable, participants were categorized as having depression if they answer “Yes” to the question “(Ever told) you have a depressive disorder (including depression, major depression, dysthymia, or minor depression)? Participants will be categorized as not having depression if they answered “No.”

The study could be affected based on the following confounders such as participant’s age, level of education, and time commitment status: *1)* age which was categorized in two groups: *1* between 25-44 years and *2* between 45-64 years; *2)* education level was categorized in 3 groups: No High School, High School and College or higher; and *3)* for time commitment status was categorized in two groups: Full-time occupation and Not Full-time occupation.

We employed four steps in our data statistical analysis: *1)* a descriptive analysis of characteristics of the sample, using frequency and percentage for qualitative variables; and measures of central tendency (mean and median) and the corresponding measure of spread (standard deviations and ranges) to described quantitative variables; *2)* bivariate analyses to determine the association between baseline characteristics/confounders and exposure (cannabis use) along with the association between characteristics/confounders and the outcome (depression disorders); *3)* collinearity assessment to determine the correlation between the confounders (age, level of education, and time commitment status) and *4)* a multivariate regression analysis using logistic regression to determine the crude and adjusted association between the exposure and outcome (crude and adjusted, respectively). We considered a p-value ≤ 0.05 as statistically significant. SPSS software was used for these analyses.

## Ethical aspects

As this is a cross-sectional study conducted in 2022, there is minimal risk because there were no vulnerable populations involved since we will be using results from the 2018 BRFSS database. Because we are not doing a secondary analysis of data, we will not have to face the ethical considerations that are normally associated with data collection first-hand, as we will not have to directly interact with human subjects. Despite not having contact with people, clinical practitioners may benefit from our research since we probably portray the negative/positive aspects of using cannabis. It may also be resourceful due to the overarching legalization happening globally. The reverse causality was investigated via our research, and it provides further insight into the use of cannabis as treatment.

## Results

The sample included 57,757 individuals between the ages of 25 and 64 years. *Tables 1* and *2* show the baseline characteristics of respondents between ages 25 and 64 years and its relationship with cannabis use and depression, respectively. *Table 3* shows the unadjusted and adjusted binary logistic regressions for potential confounders.

**Table 1.** Baseline Characteristics of American Adults Between Ages 25-64 years who responded to the 2018 BRFSS Survey by Cannabis Use in The Past 30 Days.

|  | Cannabis Use |  |  |  | p-value* |
| --- | --- | --- | --- | --- | --- |
|  | Yes<br>(N) | % | No<br>(N) | % |  |
| **Age Groups (years)** |  |  |  |  | < 0.001 |
| 25-44 | 301 | 62.8 | 4,995 | 47.9 |  |
| 45-64 | 297 | 37.2 | 9,275 | 52.1 |  |
| **Education Level** |  |  |  |  | 0.88 |
| No High School | 16 | 7.0 | 357 | 8.0 |  |
| High School | 106 | 16.7 | 2,158 | 17.0 |  |
| College or Higher | 476 | 76.3 | 11,732 | 75.1 |  |
| **Time Commitment Status** |  |  |  |  | 0.051 |
| Full-Time | 485 | 84.5 | 12,376 | 88.9 |  |
| Not Full-Time | 111 | 15.5 | 1,822 | 11.1 |  |

Individuals with a full-time occupation/schedule which includes: employed for wages, self-employed, as well as homemakers and students. Those who are categorized in the not full-time section include: individuals who have been out of work for 1 year or more, out of work for less than 1 year, retired, and unable to work.
\*Chi-square test

**Table 2.** Baseline Characteristics of American Adults Between Ages 25-64 years Who Responded to the 2018 BRFSS Survey by Depressive Disorder.

|  | Depression |  |  |  | p-value* |
| --- | --- | --- | --- | --- | --- |
|  | Yes<br>(N) | % | No<br>(N) | % |  |
| **Cannabis use** |  |  |  |  | < 0.001 |
| Cannabis Use | 142 | 21.7 | 454 | 78.3 |  |
| Non-Cannabis Use | 1,948 | 12.5 | 12,295 | 87.5 |  |
| **Age Groups (years)** |  |  |  |  | 0.004 |
| 25-44 | 3,330 | 14.4 | 18,663 | 85.6 |  |
| 45-64 | 5,026 | 12.9 | 30,682 | 87.1 |  |
| **Education Level** |  |  |  |  | 0.122 |
| No High School | 210 | 13.1 | 1,062 | 86.9 |  |
| High School | 1,167 | 12.3 | 7,156 | 87.9 |  |
| College or Higher | 6,972 | 14.1 | 40,901 | 85.9 |  |
| **Time Commitment Status<sup>a</sup>** |  |  |  |  | < 0.001 |
| Full-Time | 6,523 | 12.5 | 43,410 | 87.5 |  |
| Not Full-Time | 1,791 | 23.9 | 5,510 | 76.1 |  |

<sup>a</sup>Individuals with a full-time occupation/schedule which includes: employed for wages, self-employed, as well as homemakers and students. Those who are categorized in the not full-time section include: individuals who have been out of work for 1 year or more, out of work for less than 1 year, retired, and unable to work.
\*Chi-square test

**Table 3.**
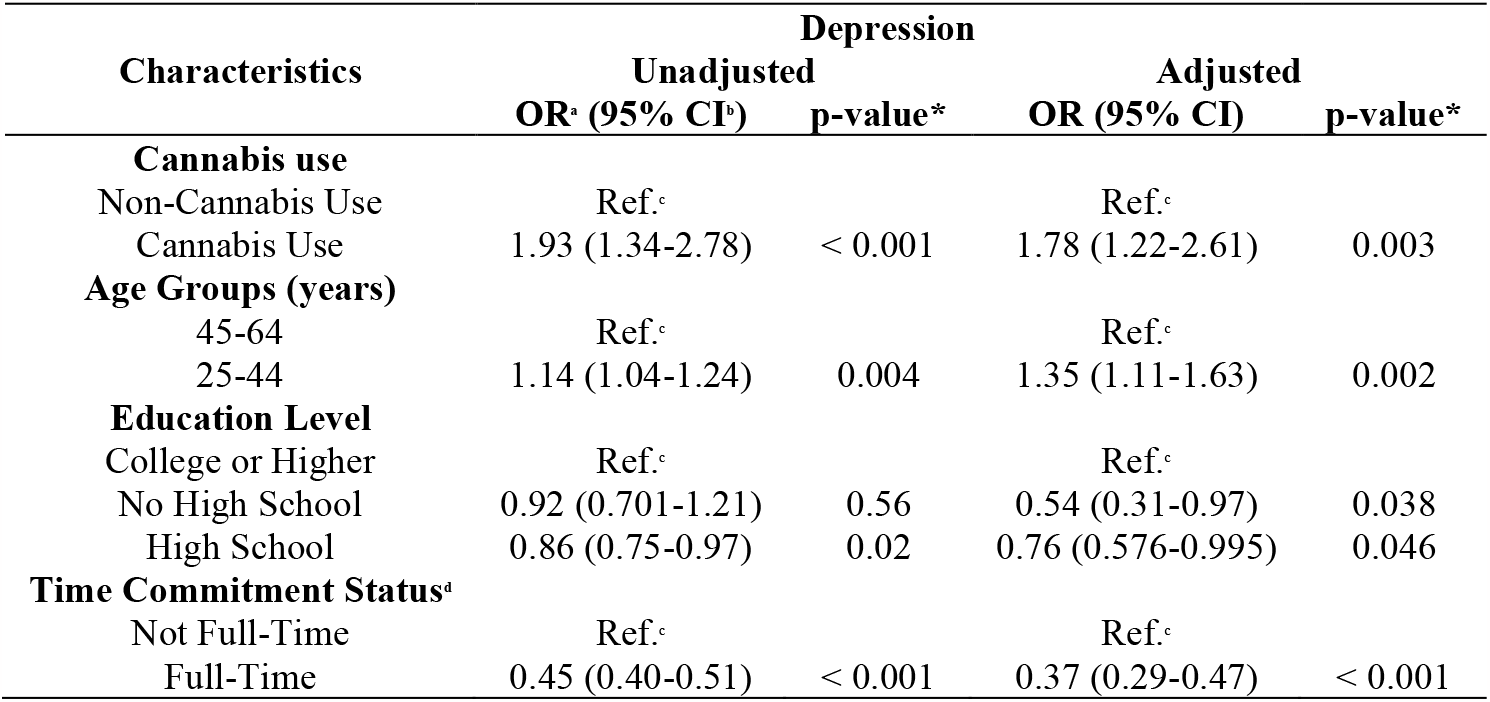
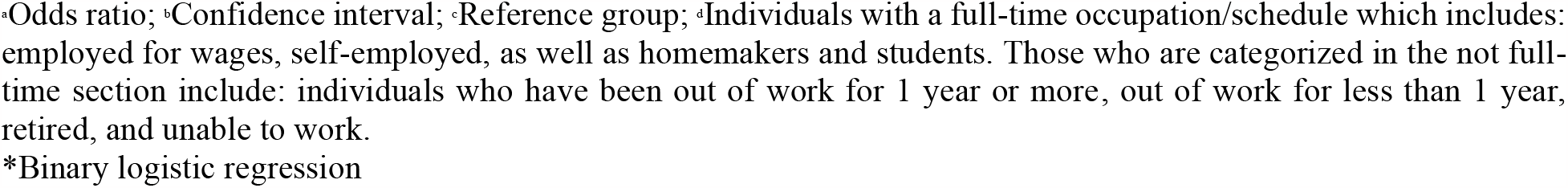
Unadjusted and Adjusted Associations Between Having Ever Been Told They Have Had a Depressive Disorder and the Use of Cannabis in the Past 30 Days.

| Characteristics | Depression |  |  |  |
| --- | --- | --- | --- | --- |
|  | Unadjusted<br>OR<sup>c</sup> (95% CI)<sup>b</sup> | p-value* | Adjusted<br>OR (95% CI) | p-value* |
| **Cannabis use** |  |  |  |  |
| Non-Cannabis Use | Ref.<sup>c</sup> |  | Ref.<sup>c</sup> |  |
| Cannabis Use | 1.93 (1.34-2.78) | < 0.001 | 1.78 (1.22-2.61) | 0.003 |
| **Age Groups (years)** |  |  |  |  |
| 45-64 | Ref.<sup>c</sup> |  | Ref.<sup>c</sup> |  |
| 25-44 | 1.14 (1.04-1.24) | 0.004 | 1.35 (1.11-1.63) | 0.002 |
| **Education Level** |  |  |  |  |
| College or Higher | Ref.<sup>c</sup> |  | Ref.<sup>c</sup> |  |
| No High School | 0.92 (0.701-1.21) | 0.56 | 0.54 (0.31-0.97) | 0.038 |
| High School | 0.86 (0.75-0.97) | 0.02 | 0.76 (0.576-0.995) | 0.046 |
| **Time Commitment Status<sup>a</sup>** |  |  |  |  |
| Not Full-Time | Ref.<sup>c</sup> |  | Ref.<sup>c</sup> |  |
| Full-Time | 0.45 (0.40-0.51) | < 0.001 | 0.37 (0.29-0.47) | < 0.001 |

•Odds ratio; •Confidence interval; •Reference group; •Individuals with a full-time occupation/schedule which includes: employed for wages, self-employed, as well as homemakers and students. Those who are categorized in the not full-time section include: individuals who have been out of work for 1 year or more, out of work for less than 1 year, retired, and unable to work.
\*Binary logistic regression

*Table 1* highlights a significant difference in cannabis use between different age groups. Specifically, the data indicates that a significantly higher proportion of younger adult participants reported using cannabis compared to senior adults. In the group of younger adult participants 63% admitted to using cannabis, while among senior adults, this figure was 37%. Another significant difference is observed in the time commitment status of the participants.

The data suggests that a substantial majority, roughly 85% of the participants, reported having a full-time commitment occupation and using cannabis. In contrast, the remaining 15% of participants that did not have a full-time commitment tended to use cannabis to a lesser extent. However, it’s important to note that this difference in cannabis usage between the two groups did not reach a level of statistical significance. This means that although there is a difference in cannabis consumption between the two groups, this difference is not large enough to be considered statistically significant. In other words, it’s possible that this difference could have occurred by chance or due to other factors, and it may not necessarily imply a causal relationship between full-time employment and cannabis use.

*Table 2* shows the relationship between time commitment status, cannabis use, and depression. The data reveal that among participants who did not have a full-time time commitment, approximately 24% of them reported cannabis use and were significantly more likely to experience depression. This finding suggests that individuals who have less demanding work schedules or who are not fully committed to a job may be more susceptible to using cannabis and experiencing symptoms of depression. Individuals with non-full-time commitments may face different stressors or have more flexibility in their daily routines, potentially leading to increased cannabis use as a coping mechanism or a means to alleviate stress.

*Table 3* presents the results of binary logistic regression analyses. The unadjusted analysis reveals that individuals who use cannabis are 93% more likely to receive a diagnosis of depression (OR: 1.93, 95% CI 1.34-2.78, p <0.001). As well, in the adjusted analysis, it is indicated that those who use cannabis are 78% more likely to have a diagnosis of depression (OR: 1.78, 95% CI 1.22-2.61, p=0.003).

Additionally, individuals under the age of 45 years were highly associated with being diagnosed with depression (OR: 1.35, 95% CI 1.11-1.63, p= 0.002). Nevertheless, individuals without a high school diploma (OR: 0.54 95% CI: 0.31-0.975, p=0.038) and those in full-time commitment (OR: 0.37 95% CI: 0.29-0.47, p<0.001) showed a notably less prone to being diagnosed with depression.

## Discussion

Bahorik et al. conducted a randomized trial, among patients with depression, non-medical cannabis use is more common and showed less improvement in depressive symptomatology, but it helps to reduce visits to the psychiatrist, showing that population with lower level of education has more tendency to use cannabis than those with higher level of education: 35.8% of the population with only high school level of education use cannabis, compared to 31.6% of the population with only some college degree; what’s more, low annual income has shown increased use of cannabis as well (6). However, medicinal cannabis users also had worse mental and physical health because the prescription or recommendation is to alleviate symptoms (e.g., chronic pain, epilepsy, hepatitis C, and HIV) but with the risk to acquire deficits in cognitive and physical capacity (6). These findings are consistent with patients who are referring depression onset and use cannabis, particularly heavy use, may be associated with an increased risk for developing depressive disorders with a need for further longitudinal exploration of the association between cannabis and developing depression as reported by Lev-ran et al. in a systematic review and meta-analysis (11). Nonetheless, published literature of Onaemo et al. show that data is equally consistent with cannabis use having a bidirectional effect on anxiety in such that chronically use of cannabis can possibly lead to anxiety; but individuals with acute anxiety could experience a reduction in symptoms following cannabis use; such a difference in the effect of cannabis could be due to dose-dependent interaction between delta-9-tetrahydrocannabinol, neurotransmitter system and endocannabinoid dysregulation (8). Additionally, the uncertain or unstable nature of non-full-time arrangements may contribute to feelings of uncertainty or instability, which can impact mental health and increase the likelihood of experiencing depression.

Most participants being engaged in full-time commitment status suggests that they have significant work-related responsibilities and may have limited time available for other activities or duties. The smaller subset of participants without full-time commitments consisted of individuals who were unemployed, retirees, or individuals with other non-traditional arrangements. The reasons for not having full-time commitments may vary and can influence their lifestyle, daily routines, and overall time availability. Understanding the distribution of participants across different time commitment status can provide insights into the potential impact of work-related factors on cannabis use and mental health outcomes. Factors such as stress levels, work-life balance, and access to resources for managing mental health may differ between those with full-time commitments and those without (12). These findings highlight the importance of considering employment and time commitments when examining the relationship between cannabis use and mental health outcomes. A study emphasizes the need to account for diverse socioeconomic factors and occupational contexts when designing interventions and treatment strategies related to cannabis use and mental well-being (13). Understanding the interplay between time commitment status, cannabis use, and depression can inform targeted interventions and support systems for individuals with non-full-time employment or flexible work arrangements. There is a need to highlight comprehensive mental health services that consider the unique challenges faced by individuals in different situations and provide appropriate resources and support for managing mental well-being. The contrasting usage patterns between age groups may be influenced by various factors, including generational attitudes towards cannabis, differences in access and availability, varying perceptions of its effects, and diverse reasons for consumption.

Carrà et al. conducted a cross-sectional analysis within the association between past-year cannabis use with major depressive episode (MDE) regardless of gender and age effects across different levels of cannabis use (14). There was also a weak dose-response relationship detected for adults with a minor gradient in MDE rates by increasing cannabis levels (14). Overall, these studies discussed the risks associated with a various degree of cannabis use, containing sufficient sample sizes using reliable databases to provide statistically significant data. Review of the current literature found a correlation between the use of cannabis and MDD, with heavy use of cannabis presenting as a significant risk factor (14). In addition, in the reverse correlation we found that cannabis will improve signs and symptoms of anxiety, has no effect on the symptoms of depression, and can cause deleterious effects on general health.

## Limitations

The unmeasured confounders that may affect the use of cannabis are plentiful and can significantly impact the findings, such as socio-economic status which may influence both their likelihood of using cannabis and their risk of experiencing depression. Furthermore, the study’s inability to prove the reverse causality of cannabis being used as a treatment for pre-depressed individuals poses an essential limitation. It leaves open the possibility that some participants might have turned to cannabis as a coping mechanism for their depressive symptoms, rather than cannabis use having a preventive effect on depression. Additionally, the potential underestimation of the prevalence of depression within the sample. The study may have missed individuals who were undiagnosed or did not seek help for their depression, leading to an incomplete picture of the relationship between cannabis use and depression. To address this limitation, future studies could employ more diverse recruitment methods, including community outreach and collaboration with healthcare providers, to capture a broader and more representative sample of individuals with depression. Moreover, the exclusion of individuals who used tobacco from the study population, it might have inadvertently skewed the results, as tobacco use is often correlated with cannabis use, and tobacco users may have different patterns of depression risk. The assumption that this exclusion group represents a significantly large population could be flawed, potentially leading to biased conclusions. While the study provides valuable insights into the association between cannabis use and depression, there are several unmeasured confounders and limitations that warrant caution in interpreting the results. However, they were not used in this survey. These confounders are as follows: gender, race/ethnicity, annual income, substance use disorders, chronic pain, medication and recreational drug use, nicotine use other than cigarettes, inadequate nutrition, and any other comorbidities.

To build a more robust understanding of this relationship, future research should adopt a multifaceted approach, addressing confounding variables, considering reverse causality, and employing more inclusive sampling methods. Factors such as genetic predisposition, family history of mental health conditions, childhood trauma, and exposure to other substances may play crucial roles in the cannabis-depression association. Integrating these variables into the analysis could yield a more nuanced understanding of the link between cannabis use and depression.

Moreover, exploring the potential moderating effects of different cannabis strains, consumption methods, and dosages could be crucial. Some evidence suggests that certain cannabinoids may have distinct effects on mental health, and understanding these nuances can be crucial in determining the overall impact of cannabis use on depression risk. Future investigations should strive to include a more comprehensive range of substance use behaviors to better understand the complexities of cannabis use and its relationship to depression.

## Conclusion

In conclusion, those who use cannabis (78%) are significantly more likely to have a diagnosis of depression. Age <45 years, was also significantly more likely to have a diagnosis of depression. Education without a high school diploma and having a full-time commitment were less likely to be associated with the diagnosis of depression. The insights gained from this research advocates the development of targeted preventive strategies to mitigate the potential negative effects of cannabis on mental health. Education campaigns could be designed to raise awareness about the risks of cannabis use, especially for vulnerable populations. Besides, mental health support systems could be strengthened to identify and assist individuals at risk of depression who might also be using cannabis.

As a final observation, further research is essential to unravel the complex relationship between cannabis use and depression. Employing longitudinal studies, experimental designs, and comprehensive control of confounding variables will provide a more accurate understanding of causality.

## Data Availability

All data produced in the present study are available upon request to the authors

## References

1. Bains N, Abdijadid S. Major Depressive Disorder. In: StatPearls [Internet]. Treasure Island (FL): StatPearls Publishing; 2023 [cited 2023 Sep 14]. Available from: http://www.ncbi.nlm.nih.gov/books/NBK559078/

2. Depressive disorder (depression) [Internet]. [cited 2023 Sep 19]. Available from: https://www.who.int/news-room/fact-sheets/detail/depression

3. Feingold D, Weinstein A. Cannabis and Depression. In: Murillo-Rodriguez E, Pandi-Perumal SR, Monti JM, editors. Cannabinoids and Neuropsychiatric Disorders [Internet]. Cham: Springer International Publishing; 2021 [cited 2023 Jul 18]. p. 67–80. (Advances in Experimental Medicine and Biology). Available from: 10.1007/978-3-030-57369-0_5

4. Chand SP, Arif H. Depression. In: StatPearls [Internet]. Treasure Island (FL): StatPearls Publishing; 2023 [cited 2023 Sep 20]. Available from: http://www.ncbi.nlm.nih.gov/books/NBK430847/

5. NCCIH [Internet]. [cited 2023 Jul 26]. Cannabis (Marijuana) and Cannabinoids: What You Need To Know. Available from: https://www.nccih.nih.gov/health/cannabis-marijuana-and-cannabinoids-what-you-need-to-know

6. Bahorik AL, Sterling SA, Campbell CI, Weisner C, Ramo D, Satre DD. Medical and non-medical marijuana use in depression: Longitudinal associations with suicidal ideation, everyday functioning, and psychiatry service utilization. J Affect Disord. 2018 Dec 1;241:8–14. Available from: https://www.sciencedirect.com/science/article/pii/S0165032717325478

7. National Academies of Sciences E, Division H and M, Practice B on PH and PH, Agenda C on the HE of MAER and R. Cannabis: Prevalence of Use, Regulation, and Current Policy Landscape. In: The Health Effects of Cannabis and Cannabinoids: The Current State of Evidence and Recommendations for Research [Internet]. National Academies Press (US); 2017 [cited 2023 Jul 26]. Available from: https://www.ncbi.nlm.nih.gov/books/NBK425763/

8. Onaemo VN, Fawehinmi TO, D’Arcy C. Comorbid Cannabis Use Disorder with Major Depression and Generalized Anxiety Disorder: A Systematic Review with Meta-analysis of Nationally Representative Epidemiological Surveys. J Affect Disord. 2021 Feb 15;281:467–75. Available from: https://www.sciencedirect.com/science/article/pii/S0165032720331335

9. 2018_BRFSS_English_Questionnaire-508.pdf [Internet]. [cited 2023 Sep 22]. Available from: https://www.cdc.gov/brfss/questionnaires/pdf-ques/2018_BRFSS_English_Questionnaire-508.pdf

10. CDC - BRFSS - Questionnaires [Internet]. 2022 [cited 2023 Jul 18]. Available from: https://www.cdc.gov/brfss/questionnaires/index.htm

11. The association between cannabis use and depression: a systematic review and meta-analysis of longitudinal studies | Psychological Medicine | Cambridge Core [Internet]. [cited 2023 Jul 18]. Available from: https://www.cambridge.org/core/journals/psychological-medicine/article/abs/association-between-cannabis-use-and-depression-a-systematic-review-and-metaanalysis-of-longitudinal-studies/B144B7AE5A3D973289DBDD99ADE21E58

12. Airagnes G, Lemogne C, Goldberg M, Hoertel N, Roquelaure Y, Limosin F, et al. Job exposure to the public in relation with alcohol, tobacco and cannabis use: Findings from the CONSTANCES cohort study. PLOS ONE. 2018 May 1;13(5):e0196330. Available from: 10.1371/journal.pone.0196330

13. Hamieh N, Airagnes G, Descatha A, Goldberg M, Limosin F, Roquelaure Y, et al. Atypical working hours are associated with tobacco, cannabis and alcohol use: longitudinal analyses from the CONSTANCES cohort. BMC Public Health. 2022 Sep 29;22(1):1834. Available from: 10.1186/s12889-022-14246-x

14. Trends of Major Depressive Episode among People with Cannabis Use: Findings from the National Survey on Drug Use and Health 2006–2015 - Giuseppe Carrà, Francesco Bartoli, Cristina Crocamo, 2019 [Internet]. [cited 2023 Jul 18]. Available from: https://journals.sagepub.com/doi/10.1080/08897077.2018.1550464?url_ver=Z39.88-2003&rfr_id=ori:rid:crossref.org&rfr_dat=cr_pub%20%200pubmed

